# Risk Stratification of Advanced Fibrosis in HIV Patients With Hepatic Steatosis Using the NAFLD Fibrosis and BARD Scores

**DOI:** 10.1101/2023.07.07.23292294

**Authors:** George A. Yendewa, Ana Khazan, Jeffrey M. Jacobson

**Affiliations:** Department of Medicine, Case Western Reserve University School of Medicine, Cleveland, Ohio, USA; Division of Infectious Diseases and HIV Medicine, University Hospitals Cleveland Medical Center, Cleveland, Ohio, USA

**Keywords:** HIV, liver, steatosis, obesity, NAFLD, NASH, metabolic disorders, diagnostics

## Abstract

**Background:** Nonalcoholic fatty liver disease (NAFLD) is increasingly prevalent in people with HIV (PWH), yet the risk factors for disease progression are poorly understood, due to inadequate surveillance. We employed non-invasive methods to estimate the prevalence and associated factors of advanced NAFLD in PWH.

**Methods:** We conducted a retrospective study of PWH enrolled in our clinic from 2005 to 2022. We employed imaging (ultrasound, computer tomography, magnetic resonance imaging, and transient elastography) or biopsy reports to identify cases of hepatic steatosis. We excluded patients with harmful alcohol use, hepatitis B or C infection, and other specified etiologies. We used the NAFLD Fibrosis Score (NFS), BARD Score, AST to Platelet Index (APRI), and Fibrosis-4 (FIB-4) Score to stratify fibrosis. We used logistic regression to identify predictors of advanced fibrosis.

**Results:** Among 3959 PWH in care, 1201 had available imaging or liver biopsies. After exclusions, 114 of the remaining 783 had evidence of hepatic steatosis (prevalence 14.6%). The majority were male (71.1%), with mean age 46.1 years, and mean body mass index (BMI) 31.4 ± 8.1 kg/m^2^. About 24% had lean NAFLD (BMI < 25 kg/m^2^). Based on the NFS, 27.2% had advanced fibrosis, which was corroborated by estimates from the other scores. In adjusted regression analysis, advanced fibrosis was associated with BMI > 35 kg/m^2^ (4.43, 1.27-15.48), thrombocytopenia (4.85, 1.27-18.62) and hypoalbuminemia (9.01, 2.39-33.91).

**Conclusion:** We found a NAFLD prevalence of 14.6%, with 27.2% of cases having advanced fibrosis. Our study provides practical insights into the surveillance of NAFLD in PWH.

## INTRODUCTION

Non-alcoholic fatty liver disease (NAFLD) is a growing global health problem. NAFLD results from the accumulation of fat in hepatocytes (steatosis) in the absence of a specified etiology such as alcohol use disorder, hepatitis B (HBV) or C (HCV), autoimmune hepatitis, Wilson’s disease, and other hereditary metabolic disorders [1]. Clinically, two main phenotypes are recognized: non-alcoholic fatty liver (NAFL), which is characterized by the accumulation of fat in hepatocytes with minimal inflammation/damage, and non-alcoholic steatohepatitis (NASH), in which significant hepatocellular damage and/or fibrosis has occurred [1]. According to recent estimates, NALFD affects about 37.8% of the general population worldwide, a dramatic rise from a prevalence of 25.5% before 2005 [2]. The growing problem of NALFD/NASH closely parallels rising trends in the global burden of classic risk factors such as obesity, type 2 diabetes mellitus, and the metabolic syndrome [1]. The presence of the NASH phenotype is associated with a higher risk of progression to cirrhosis and hepatocellular carcinoma (HCC) [3]. Consequently, NAFLD/NASH has now surpassed HCV as the leading indication for liver transplantation in developed countries [4].

NAFLD is a frequent, yet underappreciated cause of chronic liver disease among people living with HIV (PWH), however, prevalence estimates may vary widely depending on the diagnostic method used. A systematic review and meta-analysis of 10 studies published in 2017 estimated a NALFD prevalence of 35% among HIV mono-infected individuals [5], which is comparable to NAFLD rates reported in the general population [2]. However, in addition to established risk factors for NAFLD [1], PWH are susceptible to multiple pathophysiologic mechanisms of hepatocellular injury which may further increase their risk of NAFLD. Specifically, HIV-associated inflammation and immune activation accompanying HIV replication in Kupfer and hepatic stellate cells induces mitochondrial toxicity and promotes a profibrogenic microenvironment within the liver parenchyma [6–8]. Additional HIV-specific mechanisms that have been implicated in the development of NAFLD include HIV-induced enteropathy and alterations in the gut microbiome [7, 9, 10] and toxicities associated with antiretroviral treatment (ART) regimens such as older generation nucleos(t)ide reverse transcriptase inhibitors (NTRIs), protease inhibitors (PIs), and the newer generation integrase strand inhibitors (INSTI) [11–13].

As the life expectancy of PWH has dramatically increased in the era of combination ART, it is anticipated that PWH will remain at increased risk of NAFLD and other noncommunicable diseases associated with aging [14, 15]. Unfortunately, current management strategies for NAFLD such as lifestyle modifications and pharmacologic therapy have limited efficacy in advanced disease, which often requires more extreme interventions including bariatric surgery or liver transplantation [14, 15]. It is therefore essential that NAFLD is identified early for appropriate counseling, risk stratification, and management. However, diagnosing NAFLD can be challenging even in well-resourced settings. Liver biopsy, which is the gold standard for diagnosis and staging, is invasive and carries inherent risks. Noninvasive imaging methods such as transient elastography have been validated in PWH and provide highly accurate and reliable measurements of liver fibrosis [16–18]; however, they are costly and not always available in routine clinical practice. As a result, several predictive non-invasive scoring systems have been developed such as the NAFLD Fibrosis Score (NFS) [19] and the BARD Score [20]. These scores utilize routine laboratory tests and anthropometric measures and have demonstrated acceptable diagnostic performance, which has led to their increasing use as alternative diagnostic tools for detecting NAFLD in both the general population and PWH [19–22].

In this study, we aimed to estimate the prevalence of NAFLD and identify risk factors associated with advanced disease (i.e., NASH) in a university hospital-based HIV cohort in Northeast Ohio in the United States using non-invasive scores for detecting liver fibrosis.

## METHODS

### Study Design, Setting and Cohort

We conducted a retrospective cross-sectional study by reviewing the medical records of HIV-infected patients who received routine care at the Special Immunology Unit (SIU) at the University Hospitals Cleveland Medical Center in Cleveland, Ohio, United States from 2005 to 2022. The SIU was established in 1985 to provide comprehensive clinical care to PWH or those at risk of acquiring HIV infection, including HIV testing, counseling and support services, ART, pre-exposure prophylaxis, and clinical trials. To date, the SIU has provided clinical care to over 4000 PWH, with 1214 patients actively in care in 2022.

### Data Extraction and Study Definitions

The study inclusion criteria were age ≥ 18 years with at least one visit to the SIU between 2005 and 2022, documented evidence of HIV infection (i.e., antibody/antigen testing, polymerase chain reaction, or Western blot), and evidence of hepatic steatosis based on characteristic findings on ultrasonography, computed tomography, magnetic resonance imaging, or transient elastography, or liver biopsy report. Exclusion criteria were documented significant alcohol use disorder, HBV or HCV infection, autoimmune hepatitis, Wilson’s disease, hemochromatosis, primary biliary sclerosis, drug-induced liver injury, and other liver diseases of specified etiology such as cholestatic, metabolic, or genetic liver-related diseases.

We extracted demographic, clinical, and laboratory data from the medical records of patients who met the study inclusion criteria. We collected data on age, sex, race/ethnicity, body mass index (BMI), alcohol use, smoking history, drug history, comorbidities (i.e., hypertension, prediabetes, diabetes, obesity, and dyslipidemia), angiotensin-converting enzyme inhibitors (ACE-Is) and angiotensin receptor blockers (ARBs), statins, anti-diabetic agents (i.e., insulin and metformin), ART history and HIV indices (i.e., CD4 count and viral load). Additional laboratory data collected included platelet count, hemoglobin A1c (HbA1c), serum aspartate transaminase (AST), alanine transaminase (ALT), bilirubin (total and direct), albumin, and lipid panel (total cholesterol, high-density lipoprotein (HDL-C), low-density lipoprotein (LDL-C), and triglycerides(TG). All laboratory data extracted were within 6 months of the diagnosis of hepatic steatosis.

We calculated the BMI using the formula mass(kg)/height^2^(m^2^) and classified participants as underweight (i.e., BMI < 18.5 kg/m^2^), normal (i.e., BMI 18.5-24.9 kg/m^2^), overweight (i.e., BMI 25-29.9 kg/m^2^), obese (i.e., BMI 30-34.9 kg/m^2^), and morbidly obese (i.e., BMI ≥ 35 kg/m^2^) [23]. We defined prediabetes as an HbA1c of 5.7% to 6.4%, and diabetes as an HbA1c ≥ 6.5% or being on anti-diabetic treatment [24]. We defined hypertension as documented blood pressure ≥ 140/90 mmHg or being on antihypertensive treatment [25]. We used the harmonized criteria for the metabolic syndrome to define dyslipidemia thresholds as follows: hypertriglyceridemia (TG ≥ 150 mg/dL), low HDL-C (i.e., HDL-C < 40 mg/dL for males or < 50 mg/dL for females), high LDL-C (i.e., LDL-C > 100 mg/dL), and hypercholesterolemia (i.e., total cholesterol ≥ 200 mg/dL) [26].

### Non-invasive Assessments of Liver Fibrosis

We used four validated non-invasive scoring systems to estimate the degree of liver fibrosis. The Aspartate Transaminase to Platelet Ratio (APRI) Score was calculated using the formula:

APRI = [AST (IU/L)/AST (Upper Limit of Normal) (IU/L)]/[Platelet Count (109/L)] × 100 [27]. The following thresholds defined the degree of fibrosis: APRI < 0.5 for normal liver histology, APRI 0.5–1.5 for significant fibrosis, and APRI > 1.5 for advanced fibrosis [27].

We calculated the FIB-4 Score using the formula:

FIB-4 = [Age (years) × AST (IU/L)]/[Platelet Count (10^9^/L) × [ALT (IU/L)]^1/2^] [28]. A FIB-4 score < 1.45 was classified as normal liver, a FIB-4 Score of 1.45 to 3.25 as significant fibrosis, and a FIB-4 Score > 3.25 as advanced fibrosis [28].

We calculated the BARD Score using the BMI, AST/ALT ratio, and the presence or absence of type 2 diabetes mellitus to predict advanced fibrosis in NAFLD as originally described by Harrison *et al* [20]. The maximum possible BARD Score is 4, with BMI > 28 kg/m^2^ scored as 1, AS/ALT > 0.8 scored as 2 and DM scored as 1. A BARD Score of 0-1 indicates a low probability of fibrosis and can rule out the need for a liver biopsy, while a score of 2-4 indicates an odds for advanced fibrosis of 17 (95% CI 9.2 to 31.9), with a negative predictive value of 96% [20].

Lastly, we calculated the NFS using the formula:

NFS = −1.675 + 0.037 × Age (year) + 0.094 × BMI (kg/m^2^) + 1.13 × IFG/Diabetes (Yes = 1, No = 0) + 0.99 × AST/ALT - 0.013 × Platelet Count (×10^9^/L) - 0.66 × Albumin (g/dL) [19], where IFG represents the presence of impaired fasting glucose. The degree of fibrosis was categorized as follows: NFS < −1.5 for normal liver, NFS ≥ −1.5 to < 0.67 for mild/moderate fibrosis, and NFS ≥ 0.67 for a high probability of advanced fibrosis or NASH [19].

### Statistical Analyses

We performed statistical analyses using SPSS Version 29.0 (Armonk, NY; IBM Corp). We reported categorical variables as frequencies (percentages) and assessed associations using Pearson’s chi-square or Fisher’s exact tests. We presented continuous variables as means (standard deviations, SD) and assessed associations using the independent Student *t*-test. We used logistic regression models to identify associations between advanced and mild/moderate NAFLD. We evaluated known risk factors and potential disease modifiers of NAFLD in univariate analysis. Only covariates which attained a p-value of < 0.20 in the univariate analysis were included in the multivariate regression model. We reported associations as crude (OR) and adjusted odds ratios (AOR) with 95% confidence intervals (CI). In all analyses, p < 0.05 was considered statistically significant.

### Ethical Approval

We obtained ethical approval from the University Hospitals Cleveland Medical Center Institutional Review Board. Informed consent was not deemed necessary since the study involved only de-identified information.

## RESULTS

### Prevalence of NAFLD

Among 3959 PWH who received care at our clinic between 2005 and 2022, 1201 had available imaging or liver biopsy reports. A further 418 PWH with known etiologies for liver disease were excluded. Of the remaining 783, 114 had imaging or biopsy evidence of hepatic steatosis, yielding a NAFLD prevalence of 14.6% (Figure 1).

**Figure 1.**
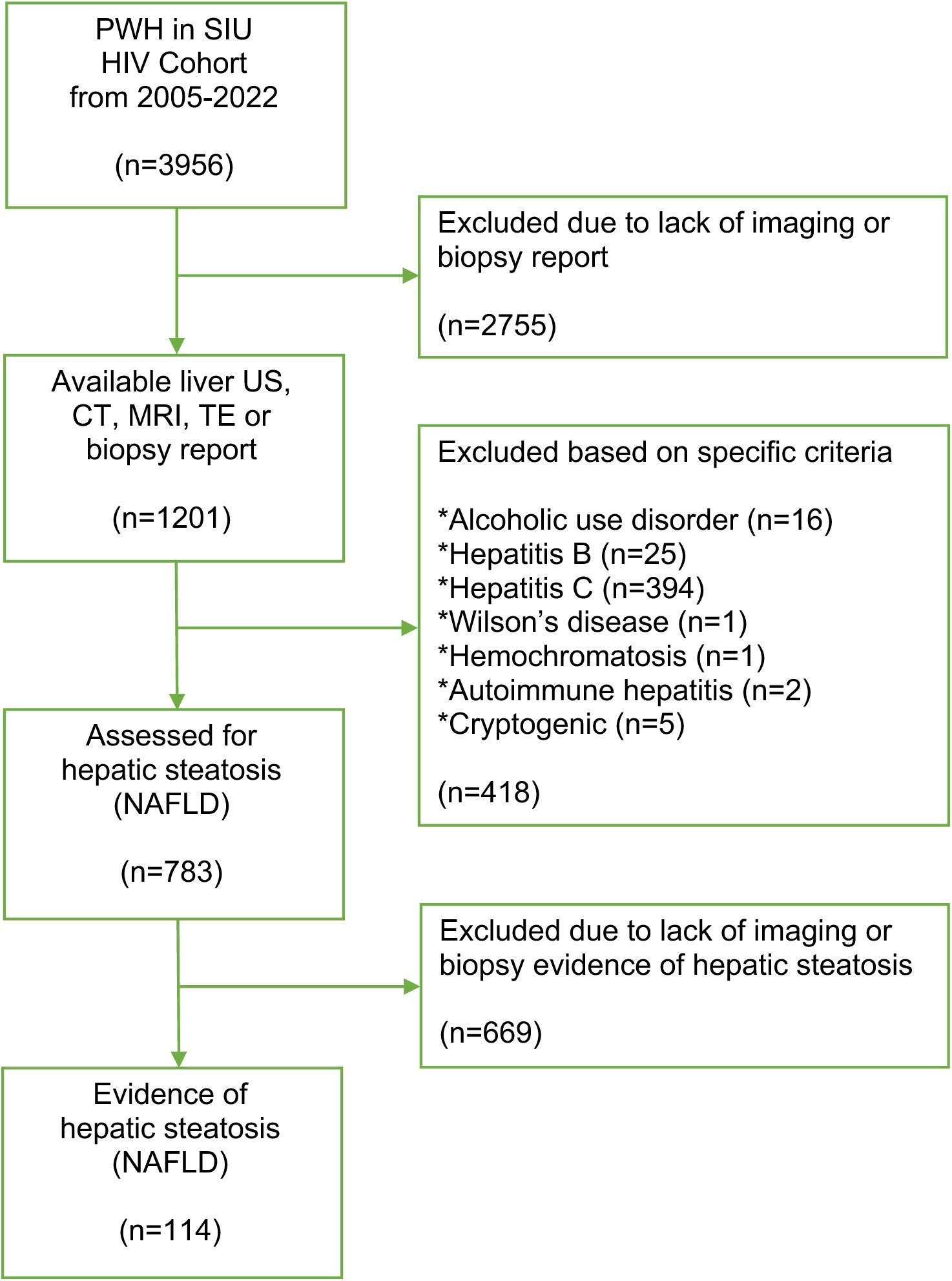
Flow chart demonstrating inclusion and exclusion of study participants. CT, computer tomography; HIV, human immunodeficiency virus; MRI, magnetic resonance imaging; NAFLD, nonalcoholic fatty liver disease; PWH, people with HIV; SIU, Special Immunology Unit; TE, transient elastography. *Cases are not necessarily mutually exclusive to other conditions.

### General characteristics and classification of NAFLD

Table 1 describes the characteristics of the study population. Of the 114 PWH with NAFLD, 81 (71.1%) were male. The mean age was 46.1 ± 11.0 years and the majority (59, 51.8%) were Black. The mean BMI was 31.4 ± 8.1 kg/m^2^, with 24 (21.1%) categorized as normal/underweight (BMI < 25 kg/m^2^), while 35 (30.7%) were morbidly obese (BMI > 35 kg/m^2^). The majority were current or past smokers (73, 64.0%) and about one-third (36, 31.6%) had a history of drug use. The majority had hypertension (65, 57.0%), over one-third (42, 36.8%) were diabetic, and 25 (21.9%) had hyperlipidemia. The mean CD4 count was 574 ± 418 cells/mm^3^ and 107 (93.9%) were virologically suppressed. All were on ART, with 96 (84.2%) on a tenofovir-based regimen, 55 (48.2%) on INSTIs, 33 (28.9%) on PIs, and 26 (22.8%) on NNRTIs.

**Table 1.** Baseline demographic and clinical characteristics of PWH with NAFLD.

| Characteristics | N (%) |
| --- | --- |
| Male sex | 81 (71.1) |
| Age (years) |  |
| Mean ± SD | 46.1 ± 11.0 |
| $\leq 34$ | 19 (16.7) |
| 35-44 | 25 (21.9) |
| 45-54 | 44 (38.6) |
| $\geq 55$ | 26 (22.8) |
| Race/ethnicity |  |
| Black | 59 (51.8) |
| White | 51 (44.7) |
| Hispanic | 3 (2.6) |
| Asian | 1 (0.9) |
| Body mass index (kg/m<sup>2</sup>) |  |
| Mean ± SD | 31.4 ± 8.1 |
| <18.5 (underweight) | 3 (2.6) |
| 18.5-24.9 (normal) | 21 (18.4) |
| 25.0-29.9 (preobesity) | 32 (28.1) |
| 30.0-34.9 (obesity) | 23 (20.2) |
| $\geq 35.0$ (morbid obesity) | 35 (30.7) |
| Life-style associated risk factors |  |
| Smoking | 73 (64.0) |
| Drug use | 36 (31.6) |
| Comorbidities |  |
| Hypertension | 65 (57.0) |
| Hyperlipidemia | 25 (21.9) |
| Diabetes | 42 (36.8) |
| CD4 count (cells/mm<sup>3</sup>) |  |
| Mean ± SD | 574 ± 418 |
| $\geq 200$ | 94 (82.5) |
| HIV RNA < 20 copies/mL | 107 (93.9) |
| ART regimen |  |
| Tenofovir-based | 96 (84.2) |
| INSTI-based | 55 (48.2) |
| PI-based | 33 (28.9) |
| NNRTI-based | 26 (22.8) |
| Antihypertensives |  |
| ACE-I or ARB | 41 (36.0) |
| Others | 24 (21.1) |
| Lipid-lowering agent |  |
| Statin | 47 (41.2) |
| Others | 6 (5.3) |
| Antidiabetic treatment |  |
| Insulin plus other(s) | 16 (14.0) |
| Insulin alone | 8 (7.0) |
| Metformin | 12 (10.5) |
| APRI Score |  |
| Mean ± SD | 1.75 ± 4.01 |
| > 1.5 (F3-F4) | 36 (29.3) |
| FIB-4 Score |  |
| Mean ± SD | 3.75 ± 6.96 |
| > 3.25 (F3-F4) | 28 (24.6) |
| BARD Score |  |
| Mean ± SD | 2.32 ± 1.08 |
| 0-1 | 25 (21.9) |
| 2-4 | 89 (78.1) |
| NAFLD Score |  |
| Mean ± SD | -0.09 ± 3.02 |
| < -1.45 (F0) | 36 (31.6) |
| -1.45 to 0.67 (F1-F2) | 47 (41.2) |
| ≥ 0.67 (F3-F4) | 31 (27.2) |

ACE-I, angiotensin-converting enzyme inhibitor; APRI, aspartate aminotransferase (AST) to platelet ratio index; ARB, angiotensin receptor blocker; ART, antiretroviral therapy; BARD score, body mass index, AST/alanine transaminase (ALT) ratio and diabetes score; FIB-4, fibrosis index based on 4 factors; HDL-C, high density lipoprotein; INSTI, integrase strand transfer inhibitor; LDL-C, low density lipoprotein; NAFLD, nonalcoholic fatty liver disease; NNRTI, non-nucleoside reverse transcriptase inhibitors; PI, protease inhibitors.

Based on the NFS estimates, 36 (31.6%) had normal liver architecture (NFS < −1.5), 47 (41.2%) had mild-to-moderate fibrosis (−1.5 ≤ NFS < 0.67), and 31 (27.2%) had advanced fibrosis/NASH (NFS ≥ 0.67). These results were in agreement with estimates from the APRI, FIB-4, and BARD scores (Table 1). According to the BARD scores, 25 patients (21.9%) had a low probability of fibrosis and could avoid having a liver biopsy.

### Comparison of patient characteristics of mild/moderate and advanced fibrosis

Compared with those with mild/moderate fibrosis, PWH with advanced fibrosis/NASH were significantly more likely to have higher BMI (mean 35.6 ± 9.8 kg/m^2^ vs 29.8 ± 6.9 kg/m^2^, p < 0.001), lower platelet counts (mean 171 ± 80 x10^9^/L vs 241 ± 95 x10^9^/L, p < 0.001), elevated AST (mean 198 ± 280 IU/L vs 71 ± 79 IU/L, p < 0.001), higher direct bilirubin levels (mean 1.0 ± 1.7 mg/mL vs 0.5 ± 0.9 mg/mL, p = 0.027), and lower albumin levels (mean 3.1 ± 0.9 mg/dL vs 4.0 ± 0.7 mg/dL, p < 0.001) (Table 2). There was no difference between the groups in terms of sociodemographic factors, comorbidities, CD4 count, HIV viremia, class of ART, serum lipid profile, or potential modifiers of liver fibrosis such as antihypertensives, antidiabetics, and statins.

**Table 2.** Comparison of mild/moderate and advanced fibrosis by selected parameters.

| Characteristics | Advanced fibrosis<br>F3-F4<br>(n=31) | Mild/moderate<br>fibrosis F0-F2<br>(n=83) | p-Value |
| --- | --- | --- | --- |
| Age (years)<sup>a</sup> | 46.4 ± 11.4 | 46.0 ± 10.9 | 0.884 |
| Male sex, n (%) | 18 (58.1) | 63 (75.9) | 0.062 |
| Race/ethnicity, n (%) |  |  |  |
| Black | 17 (54.8) | 42 (50.6) | 0.841 |
| White | 14 (45.2) | 37 (44.6) |  |
| Hispanic | - | 3 (3.6) |  |
| Asian | - | 1 (1.2) |  |
| Body mass index (kg/m<sup>2</sup>) | 35.6 ± 9.8 | 29.8 ± 6.9 | **<0.001** |
| Smoking, n (%) | 17 (54.8) | 56 (67.5) | 0.211 |
| Drug use, n (%) | 6 (19.4) | 30 (36.1) | 0.086 |
| Hypertension, n (%) | 14 (45.2) | 51 (61.4) | 0.118 |
| Prediabetes or diabetes, n (%) | 14 (45.2) | 39 (47.0) | 0.862 |
| CD4 count (cells/mm<sup>3</sup>) | 470 ± 335 | 613 ± 440 | 0.106 |
| HIV RNA < 20 copies/mL, n (%) | 1 (3.2) | 6 (7.2) | 0.428 |
| ART regimen, n (%) |  |  |  |
| Tenofovir-based | 27 (87.1) | 69 (83.1) | 0.606 |
| NNRI-based | 8 (25.8) | 18 (21.7) | 0.855 |
| PI-based | 8 (25.8) | 25 (30.1) |  |
| INSTI-based | 15 (48.4) | 40 (48.2) |  |
| ACE-I or ARB use | 12 (38.7) | 29 (34.9) | 0.709 |
| Statin use, n (%) | 9 (29.0) | 38 (45.8) | 0.106 |
| Insulin, n (%) | 2 (6.5) | 6 (7.2) | 0.885 |
| Metformin, n (%) | 3 (9.7) | 17 (20.5) | 0.177 |
| Platelets (x10<sup>9</sup>/L)<sup>a</sup> | 171 ± 80 | 241 ± 95 | **<0.001** |
| AST (IU/L)<sup>a</sup> | 198 ± 280 | 71 ± 79 | **<0.001** |
| ALT (IU/L)<sup>a</sup> | 59 ± 55 | 76 ± 73 | 0.234 |
| Total bilirubin (mg/dL)<sup>a</sup> | 1.8 ± 2.7 | 1.2 ± 1.6 | 0.170 |
| Direct bilirubin (mg/dL)<sup>a</sup> | 1.0 ± 1.7 | 0.5 ± 0.9 | **0.027** |
| Albumin (g/dL)<sup>a</sup> | 3.1 ± 0.9 | 4.0 ± 0.7 | **<0.001** |
| Total cholesterol (mg/dL)<sup>a</sup> | 177 ± 56 | 188 ± 47 | 0.302 |
| HDL-C (mg/dL)<sup>a</sup> | 44 ± 23 | 43 ± 18 | 0.929 |
| LDL-C (mg/dL)<sup>a</sup> | 102 ± 65 | 102 ± 45 | 0.992 |
| Triglycerides (mg/dL) | 265 ± 190 | 235 ± 167 | 0.410 |
| APRI<sup>a</sup> | 4.6 ± 8.3 | 1.1 ± 1.5 | **<0.001** |
| FIB-4<sup>a</sup> | 8.8 ± 11.6 | 1.9 ± 2.0 | **<0.001** |
| BARD score<sup>a</sup> | 3.0 ± 0.7 | 2.1 ± 1.1 | **<0.001** |

ACE-I, angiotensin-converting enzyme inhibitor; APRI, aspartate aminotransferase (AST) to platelet ratio index; ARB, angiotensin receptor blocker; ART, antiretroviral therapy; BARD score, body mass index, AST/alanine transaminase (ALT) ratio and diabetes score; FIB-4, fibrosis index based on 4 factors; HIV, human immunodeficiency virus; INSTI, integrase strand transfer inhibitor; HDL-C, high density lipoprotein; LDL-C, low density lipoprotein; NAFLD, nonalcoholic fatty liver disease; NNRTI, non-nucleoside reverse transcriptase inhibitors; PI, protease inhibitors; RNA, ribonucleic acid.
<sup>a</sup> mean ± standard deviation

### Predictors of advanced fibrosis

In adjusted multiple logistic regression analysis, advanced fibrosis was significantly associated with BMI > 35 kg/m^2^ (AOR 4.43, 95% CI [1.27-15.48]; p = 0.020), thrombocytopenia (AOR 4.85, 95% CI [1.27-18.62]; p = 0.021) and hypoalbuminemia (AOR 9.01, 95% CI [2.39-33.91]; p < 0.001) (Table 3).

**Table 3.**
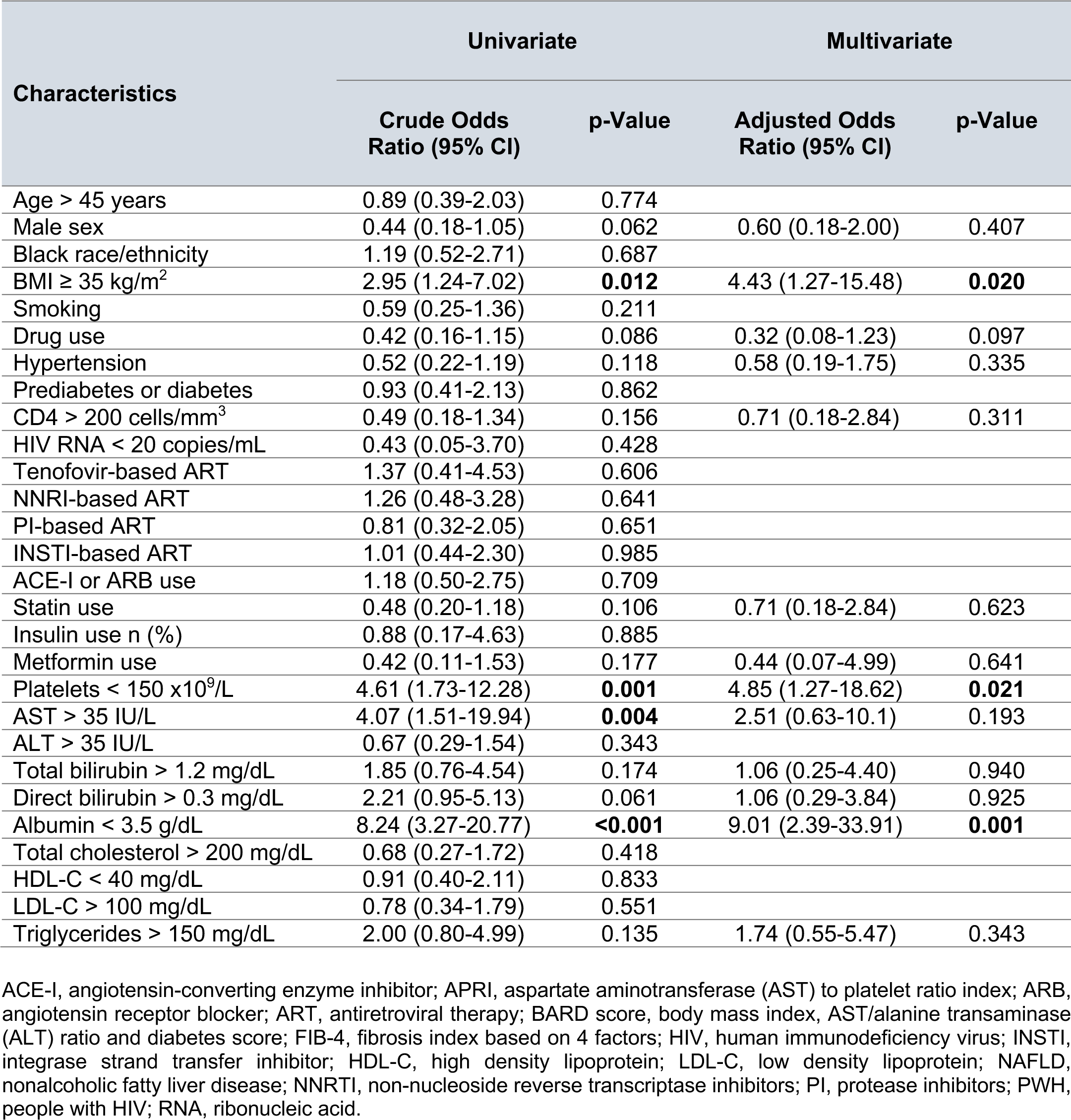
Factors Associated With Advanced NAFLD in PWH.

## DISCUSSION

In this study, we estimated NAFLD prevalence of 14.6% in our HIV cohort, of which 27.2% had advanced fibrosis or NASH using non-invasive scores. Comparatively, HIV cohort studies from North America and Europe have reported higher NAFLD prevalence rates ranging from 25% to 75% [30–34]. Several factors could account for the underdiagnosis of NAFLD among PWH. Firstly, many studies, like others, primarily rely on liver ultrasonography, readily available but less sensitive diagnostic method compared with liver biopsy and transient elastography. Secondly, classic NAFLD symptoms may be absent, subclinical, or overlap with more frequently encountered conditions such as ART-related toxicities, metabolic disorders, and co-infections with HBV or HCV. Additionally, healthcare providers may have limited awareness about NAFLD in the context of HIV; this can result in delayed identification, risk stratification, and management of NAFLD.

Our study had a few findings worthy of further discussion. Firstly, despite the well-known association between high BMI and NAFLD, a significant proportion (up to 24%) of our patients with NAFLD had normal BMI or were underweight, a condition referred to as lean NAFLD [35]. Studies have reported that between 7% and 20% of people diagnosed with NAFLD fall into this category [36–38], highlighting the complex relationship between body weight and liver health. The underlying pathophysiologic mechanisms of lean NFALD has not been elucidated; however, the emerging evidence points to distinct mechanisms contributing to NAFLD in the absence of central visceral adiposity [35]. These include impaired glucose metabolism, dysfunctional adipose tissue, and genetic factors such as carriage of the *PNPLA3* minor allele [39]. Furthermore, recent genome-wide investigations have revealed a potential association between NAFLD susceptibility and specific *HLA* alleles, such as *HLA-B*54:01*, suggesting a potential influence of gut microbiota alterations [40]. These findings highlight the need for further research to better understand the pathophysiologic mechanisms of lean NAFLD. Moreover, clinicians should be vigilant in considering the possibility of NAFLD in non-obese patients presenting with liver-related symptoms, unexplained derangements in liver function tests, or other relevant risk factors.

Secondly, all non-invasive scores used were in agreement in stratifying liver fibrosis. Approximately 25-30% of our cohort had advanced fibrosis, while about 22% had a low probability of fibrosis, effectively ruling out the necessity for a liver biopsy. The BARD Score in particular merits further consideration as it offers some advantages over the NFS, APRI, and FIB-4. The BARD Score utilizes easily obtainable clinical parameters (i.e., BMI, AST/ALT ratio, and the presence of diabetes), making it convenient for routine clinical practice [20]. In comparison, the NFS, APRI, and FIB-4 scores require more complex calculations or additional laboratory values. Furthermore, the BARD Score, like the NFS, incorporates the presence of diabetes, a key component of the metabolic syndrome and a major predictor of CVD and NAFLD risk [26]. This additional parameter enhances the predictive ability of the BARD Score in identifying advanced liver fibrosis. Moreover, extensive studies and validations [41, 42] have shown that the BARD Score, as demonstrated by Harrison et al [30] in the original study, reliably eliminates the need for liver biopsy. Taken together, the noninvasive scores offer accurate risk stratification, reduce invasive procedures, and provide cost-effective monitoring for NAFLD patients.

Thirdly, we identified surrogate markers of advanced fibrosis that could have practical implications in the routine clinical management of patients. Individuals with a BMI > 35 kg/m2 had a 4.4-fold higher odds of advanced fibrosis, while thrombocytopenia and hypoalbuminemia conferred a 4.85-fold and 9-fold increased odds of advanced liver disease, respectively. Thrombocytopenia has been widely used as a marker of advanced liver disease due to other etiologies such as cirrhosis and HCC [43, 44], as well as for progressive NALFD specifically [45, 46]. Similarly, some studies have reported associations between hypoalbuminemia and advanced NAFLD, especially in chronic kidney disease [47–49]. Furthermore, chronological changes in these indices may also have prognostic significance. For instance, a study by Kawaguchi *et al* [49] showed that a decline in serum albumin of 0.21 g/dL/year was significantly associated with a higher incidence of serious events in advanced NAFLD patients such as HCC, gastroesophageal varices, and cardiovascular events [49]. Thus, these readily obtainable indices can provide a practical means of alerting clinicians to NAFLD progression and aid in risk stratification.

Additionally, we did not observe any significant effect of statins, antihypertensives, or anti-diabetic treatment on the development of advanced fibrosis. This finding aligns with the current conflicting evidence regarding the impact of pharmacological agents on NAFLD. For example, a 2013 Cochrane review by Eslami *et al* [50] reported improvements in mortality, histological features, and biochemical profiles of NAFLD patients with statin use. However, the review included only two clinical trials with a small number of participants and a high risk of bias [51, 52]. Similarly, some studies have reported improvements in the biochemical profile, histological features, and liver-related events in NAFLD/NASH patients treated with ACE-Is/ARBs [53], insulin [54], and metformin [55], but not NASH resolution. However, recent studies have provided evidence supporting the favorable impact of semaglutide, a potent glucagon-like peptide-1 receptor agonist commonly used for the treatment of diabetes and obesity, on improving liver histology, liver function, lipid profile, or NAFLD/NASH resolution, mostly in diabetic patients [56–57], while other studies have produced contrary findings [58, 59]. Further studies are required, especially in the HIV population, to evaluate the efficacy of these pharmacological interventions in the management of NAFLD and NASH.

Lastly, we did not observe significant association between the class of ART and the degree of fibrosis, likely due to the small sample size of our study. Although ART has significantly prolonged the lifespan of PWH, it is associated with various side effects, including HIV-associated lipodystrophy and NAFLD [11–13, 35, 60]. The risk of developing NAFLD is influenced by several factors including the specific type and duration of the ART regimen, with older generation NRTIs and ritonavir-boosted PIs in particular and newer INSTIs being associated with a higher risk [60]. Oxidative stress resulting from ART-associated mitochondrial toxicity and insulin resistance appears to be the primary mechanisms contributing to the development of NAFLD [61, 62]. Despite this, the occurrence of ART-related NAFLD is relatively rare [60].

We acknowledge limitations that are inherent to our study design. These include limited generalizability due to the small sample size and restriction to a single health center. With a larger sample size, we could have potentially identified significant associations that were not apparent in our study. Furthermore, our reliance on non-invasive scores and ultrasound liver imaging may have introduced limitations and potential biases compared to more accurate diagnostic methods like liver biopsy or transient elastography. Despite these limitations, the study provides practical insights into diagnosing and managing NAFLD in PWH, which is often challenging even in well-resourced settings.

## CONCLUSION

In summary, we estimated an overall NAFLD prevalence of 14.6% in an Ohio-based HIV cohort using liver imaging criteria. Of these, we estimated an advanced fibrosis prevalence of 25% to 30% using non-invasive scores, while about 22% were stratified as low-risk for liver fibrosis, effectively reducing the need for liver biopsy. Factors associated with advanced fibrosis included BMI > 35 kg/m^2^, thrombocytopenia, and hypoalbuminemia. Our findings provide practical insights into the diagnosis and management of NALFD among PWH in routine clinical practice.

## Data Availability

All data produced in the present work are contained in the manuscript

## AUTHOR CONTRIBUTIONS

GAY and JMJ conceptualized and designed the study. GAY collected the data and conducted the statistical analysis. AK contributed important intellectual content. GAY and AK drafted the initial manuscript version. All authors critically revised and approved of the final version. GAY is acting as the guarantor of this manuscript.

## ACKLOWLEDGEMENTS

We wish to acknowledge Kimberly Robbins, Database Manager for the Case AIDS Clinical Trials Unit for her helping with data extraction and management.

## DATA AVAIL ABILITY STATEMENT

The data presented in this study are available on request from the corresponding author upon reasonable request.

